# Cancer care disruption during the COVID-19 pandemic in Ontario, Canada: A sequential mixed-methods study

**DOI:** 10.64898/2026.06.10.26355360

**Authors:** Narhari Timilshina, Danielle Jacobson, Arija Birze, Walter P Wodchis, Kerry Kuluski, Erin Strumpf, Mehdi Ammi

## Abstract

**Introduction:** The COVID-19 pandemic profoundly disrupted healthcare delivery worldwide, with cancer care among the most affected services. Prior studies documented delays in referrals, reduced specialist access, and increased provider burden. However, the extent to which these experiences were reflected at the system level remains unclear.

**Objective:** To document cancer care experiences and examine whether these experiences were reflected in population-level health system indicators across Ontario, Canada.

**Methods:** We used an exploratory sequential mixed-methods design. Qualitative data were collected through focus groups and semi-structured interviews with 32 participants, including patients with cancer (n=8), caregivers (n=5), healthcare providers (n=14), and decision-makers (n=5) across two hospital settings in Ontario, Canada. Emergent themes informed the development of quantitative indicators. We then conducted a retrospective population-based analysis of linked administrative health databases for cancer patients in Ontario (n=87,786) to assess the prevalence of identified themes.

**Results:** Four themes emerged: (I) delays in diagnosis and screening; (II) disrupted access to primary care; (III) barriers to specialist and mental health services; and (IV) fragmented care for patients with multimorbidity. Quantitative findings corroborated major themes. Screening rates declined for cervical (64.8% to 57.5%) and breast cancer (64.5% to 57.2%). While in-person primary care shifted almost entirely to virtual modalities (8.5% to 95.4%), overall visit volumes remained stable. Specialist care showed uneven patterns, with increased oncology visits but declines in cardiology and mental health services. Patients with multiple comorbidities experienced the largest reductions in non-oncology specialist care.

**Conclusion:** The pandemic disrupted key components of cancer care, particularly screening, access to certain specialist services, and care for patients with complex needs. Integrating qualitative and quantitative evidence highlights areas of system vulnerability and underscores the need for coordinated, resilient cancer care capable of maintaining essential services during future crises.

## Introduction

The COVID-19 pandemic created a global healthcare crisis that exposed and intensified pre-existing vulnerabilities in healthcare systems. During the initial waves in early 2020, access to cancer care was substantially disrupted, including screening, diagnostic procedures, specialist visits, and supportive services. Contributing factors included shifts in care delivery (e.g., reduced in-person visits), limited capacity in primary and specialty care, workforce shortages, closures of clinical spaces, and supply chain constraints affecting essential resources such as personal protective equipment, hospital beds, medications, and medical equipment (1–7). These disruptions were associated with declines across the cancer care continuum (8–13) , including a reported 39% reduction in screening, 23% decrease in diagnoses, and 28% decline in treatment during the pandemic (14). Together, these findings underscore gaps in emergency preparedness and system resilience during large-scale health crises.

Patients with chronic conditions were particularly vulnerable to these system pressures (8–13). Individuals with multimorbidity experienced greater barriers to accessing primary and specialist care, including reduced in-person visits, medication shortages, limited access to virtual services, and disruptions to preventive care (11, 15–18). Such inequities raise concerns about the uneven distribution of pandemic-related care disruptions across populations.

In Canada, similar patterns were observed, with documented declines in screening, diagnostic procedures, treatment, and follow-up services, alongside a rapid shift from in-person to virtual care (19–22). While prior research has documented these disruptions using either qualitative or population-based approaches, the two have largely been conducted separately.

To address this gap, we pursued two objectives: first, to characterize barriers to cancer care access during the COVID-19 pandemic from the perspectives of patients, caregivers, providers, and health system leaders; and second, to examine whether these themes were reflected in provincial population-level health system data. While prior studies have relied on either qualitative accounts or administrative data alone, our exploratory sequential mixed-methods design integrates both within a single analytic framework. This approach enables direct linkage between lived experiences and system-level indicators and extends qualitative insights by examining variation across subgroups, including age, comorbidity burden, immigration status, neighborhood ethnic diversity, and patient enrollment model. By combining these complementary forms of evidence, we provide a more comprehensive assessment of pandemic-related disruptions to cancer care and identify populations disproportionately affected.

## Material and Methods

### Study Design and Data Sources

This study used an exploratory sequential mixed-methods design integrating qualitative and quantitative components in Ontario, Canada (23, 24). The overall logic of the study design is described in Figure 1.

**Figure 1:**
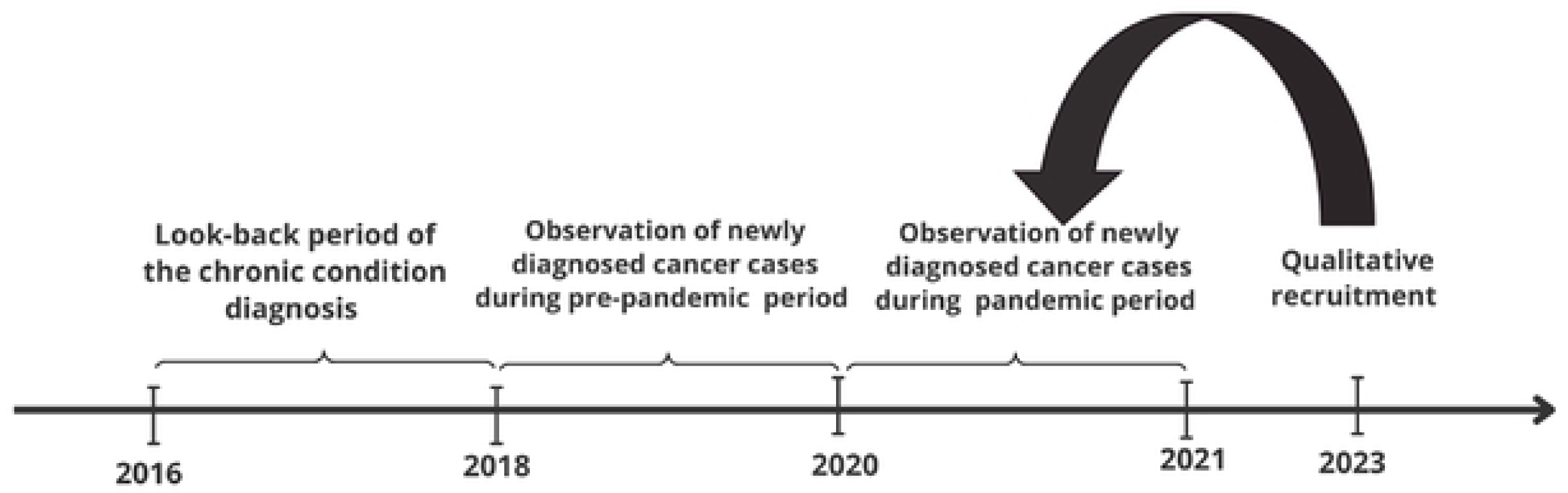
Flow chart of the study design

#### Qualitative component

The qualitative study participants were recruited between February 2023 and May 2024 from two regional cancer centres in Ontario. Eligible participants were adults (≥18 years) had received or delivered cancer care services during the COVID-19 pandemic at one of the study sites.

Data were collected through three focus groups and 26 semi-structured individual interviews, involving a total of 32 participants: patients with cancer (n=8), caregivers (n=5), healthcare providers (n=14), and healthcare system decision-makers (n=5). Interviews and focus groups were conducted via Zoom, lasted 60–90 minutes, and were audio- and video-recorded. All participants provided written informed consent and received a $25 CAD honorarium.

#### Quantitative component

The quantitative component consisted of a retrospective, population-based analysis of linked administrative health databases. The study population included adults aged 18–104 years residing in Ontario who were diagnosed with cancer (see Supplementary Table 1). Cancer cases were identified using the Ontario Cancer Registry (OCR), which captures approximately 98% of incident cancer diagnoses in the province (25).

Patient characteristics and health service utilization were obtained through linkage to multiple provincial databases, including the Ontario Health Insurance Plan (OHIP), Registered Persons Database (RPDB), Immigration, Refugees and Citizenship Canada (IRCC) database, Discharge Abstract Database (DAD), National Ambulatory Care Reporting System (NACRS), and disease-specific administrative datasets. Physician specialty information was obtained from the ICES Physician Database (26).

Comorbidities were defined using validated administrative case definitions (27–34) and included seventeen major chronic conditions such as diabetes, hypertension, chronic obstructive pulmonary disease, cardiovascular disease, mental health conditions, dementia, and rheumatoid arthritis. The final analytic cohort included n=87,786 individuals with cancer during the pre-pandemic period and n=75,417 individuals during the pandemic period.

### Data Analysis

#### Qualitative analysis

Audio recordings were transcribed verbatim, anonymized, and assigned pseudonyms. Thematic analysis was conducted using NVivo software (35, 36), following the approach described by Braun and Clarke (36, 37). Two researchers (AB and DJ) independently coded five transcripts using inductive open coding to develop an initial codebook. They met regularly to compare coding, resolve discrepancies, and refine code definitions. The codebook evolved iteratively as analysis progressed.

Themes were developed through collaborative discussion and reviewed in team meetings with another researcher (KK) to enhance analytic rigor and reflexivity. Final themes were refined and organized into core domains that informed the subsequent quantitative analyses.

#### Quantitative analysis

The quantitative component examined changes in access to cancer care using linked administrative health data. The analytic time frame was defined a priori based on data availability at the time qualitative data collection commenced in early 2023. At that point, complete update on confirmed cancer diagnosis information in administrative data were available through December 31, 2021. Given the cost and operational constraints associated with subsequent data refreshes, analyses were conducted using this fixed data cut. The pre-pandemic period was defined as January 1, 2018, to March 14, 2020, and the pandemic period as March 15, 2020, to December 31, 2021. Two cohorts of cancer patients were built based on these two periods: a cohort of patients newly diagnosed with cancer in the pre-pandemic period, and a cohort of patients newly diagnosed with cancer during the pandemic period. Variables were selected by ES, MA, and NT based on discussions with the qualitative team (AB and DJ).

Cancer screening participation was defined according to provincial program eligibility criteria (38). Colorectal cancer screening was defined as receipt of a fecal occult blood test (FOBT) or fecal immunochemical test (FIT) within the previous two years, or flexible sigmoidoscopy or colonoscopy within the previous ten years, among eligible individuals. Cervical cancer screening was defined as receipt of a Papanicolaou (Pap) test within the previous three years (or at least one cytology test within 42 months). Breast cancer screening was defined as receipt of a screening mammogram within the previous two years among eligible women. Cancer stage at diagnosis was obtained from registry data and categorized using standard staging classifications (39, 40).

Primary care utilization was measured using Ontario Health Insurance Plan (OHIP) billing claims associated with family physician, general practice, and community medicine specialty codes. Visits were classified as in-person or virtual based on billing modifiers. Both visit counts and visit modality proportions were calculated. Patient attachment in primary care status (rostered, virtually rostered, or not rostered) was determined using provincial primary care enrollment model data (41).

Specialist visits were identified using OHIP physician specialty codes, including oncology, cardiology, and endocrinology, and included both in-person and virtual encounters. Emergency department visits were identified using the National Ambulatory Care Reporting System (NACRS). Potentially avoidable visits included family practice–sensitive conditions and low-acuity visits among long-term care residents that did not require hospital admission. Outpatient mental health and addiction services were defined using validated ICD-9 and ICD-10 diagnostic algorithms linked to psychiatrist encounters or mental health–related primary care visits.

Patient-level data were structured in monthly intervals across the study period. For analysis, monthly observations were aggregated into two cohorts (pre-pandemic and pandemic), and outcomes were summarized at the patient level within each period. Continuous variables were reported as means with standard deviations or medians with interquartile ranges, as appropriate with data skewness, and categorical variables as proportions. Differences between periods were assessed using t-tests for continuous variables and chi-square tests for categorical variables.

Subgroup analyses were conducted to examine variation by sex, immigration status, neighborhood ethnic diversity, patient primary care attachment status and multimorbidity, categorized as none, one, two to four, or five or more chronic conditions. All quantitative analyses were conducted using SAS statistical software.

## Results

### Characteristics of the qualitative and quantitative study samples

#### Qualitative Sample

In total, 32 individuals participated in the qualitative portion of the study. Participants were between the ages of 30 and 79 years. Twenty-eight were female, and four were male. All participants were assigned pseudonyms to protect their anonymity.

Among patients and caregivers, educational attainment varied: seven had completed university, four had completed college, one had completed high school, and one did not report an education level. Ten identified as White, one as Black, one as European, and one did not report ethnicity. Employment status also varied: six reported working full-time, one part-time, one was self-employed, four were retired, and one did not report employment status.

Among healthcare providers and system leaders, professional experience ranged widely. Three reported up to nine years of experience; seven had between 10 and 19 years; seven had between 20 and 29 years; and two had between 30 and 39 years of professional experience. With respect to tenure at their current workplace or organization, ten reported 0–9 years, five reported 10–19 years, three reported 20–29 years, and one reported 30–39 years. All participants were assigned pseudonyms to protect anonymity.

This diverse qualitative sample provided perspectives across patient, caregiver, clinician, and leadership roles within the cancer care system.

#### Quantitative Sample

The quantitative sample included the pre-pandemic cohort of n=87,786 incident cancer patients diagnosed between January 1, 2018, and March 14, 2020, and the pandemic cohort of n=75,417 incident cancer patients diagnosed between March 15, 2020, and December 31, 2021. Descriptive statistics of these two cohorts appear in Table 1.

**Table 1:** Characteristics of the cancer patients newly diagnosed during pre-pandemic (n=87,786) and pandemic period (n=75,417) in Ontario.

| Patients characteristics |  | Cancer patients diagnosed during pre-pandemic (n=87,786) | Cancer patients diagnosed during the pandemic period (n=75,417) |
| --- | --- | --- | --- |
| Age-categories, % | 18-44 years | 8.6 | 9.7 |
|  | 45-64 years | 38.0 | 40.3 |
|  | 65-74 years | 29.1 | 29.2 |
|  | 75+ years | 24.4 | 20.8 |
| Sex, % | Female | 50.6 | 51.6 |
| Rural status, % | Yes | 12.8 | 12.6 |
| Immigration status, % | Non-immigrant | 86.8 | 86.1 |
|  | All immigrants | 11.0 | 11.6 |
|  | Refugees | 2.1 | 2.2 |
| Neighborhood ethnic diversity, % | 1 = Q1 (lowest) | 20.0 | 20.5 |
|  | 2 = Q2 | 20.4 | 21.1 |
|  | 3 = Q3 | 18.9 | 20.4 |
|  | 4 = Q4 | 19.8 | 18.6 |
|  | 5 = Q5 (highest) | 20.4 | 18.6 |
| Primary care attachment status, % | Not rostered | 2.6 | 2.4 |
|  | Rostered | 83.8 | 84.4 |
|  | Virtually rostered | 13.6 | 13.1 |
| Number of chronic conditions categories, % | None | 10.0 | 9.9 |
|  | 1 | 39.6 | 39.7 |
|  | 2 - 4 | 31.9 | 31.9 |
|  | 5+ | 18.5 | 18.5 |
| Cancer stage, % | 0 | 0.1 | 0.2 |
|  | 1 | 17.3 | 19.2 |
|  | 2 | 12.9 | 13.8 |
|  | 3 | 9.7 | 9.4 |
|  | 4 | 10.4 | 7.2 |
|  | Missing | 49.9 | 50.2 |
| Cancer type, % | Breast | 14.6 | 16.6 |
|  | Prostate | 12.4 | 13.8 |
|  | Lung | 10.9 | 8.7 |
|  | Colorectal | 9.8 | 9.8 |
|  | Lymphoma | 5.6 | 5.7 |
|  | Melanoma | 4.9 | 4.9 |
|  | Uterus | 4.2 | 4.4 |
|  | Thyroid | 4.0 | 4.6 |
|  | Kidney | 3.0 | 3.3 |
|  | Leukemia | 2.6 | 2.6 |
|  | Bladder | 2.6 | 2.6 |
|  | Oral cavity | 2.3 | 2.3 |
|  | Pancreas | 2.1 | 1.5 |
|  | Stomach | 1.8 | 1.5 |
|  | Myeloma | 1.8 | 1.7 |
|  | Ovary | 1.5 | 1.4 |
|  | Liver | 1.4 | 1.1 |
|  | Brain | 1.3 | 0.9 |
|  | Esophagus | 0.9 | 0.7 |
|  | Cervix | 0.9 | 0.9 |
|  | Testes | 0.6 | 0.7 |
|  | Larynx | 0.4 | 0.5 |
|  | Melanoma (non-cutaneous) | 0.2 | 0.2 |
|  | Other/Not otherwise specified (NOS) | 10.4 | 9.8 |

The pre-pandemic cohort had 53.5% of patients aged 65 years or older, slightly more than half were female (50.6%), and 12.8% resided in rural areas. Most patients were non-immigrants (86.8%). Patients were relatively evenly distributed across neighbourhood ethnic diversity quintiles. In the pandemic cohort, patients were slightly younger (50% aged 65 and above) and more female (51.6%), otherwise their immigration and rural residency statuses, as well as their distribution across neighbourhood by ethnic diversity were similar to the pre-pandemic cohort.

The majority were rostered to a primary care provider both in the pre-pandemic (83.8%) and pandemic cohorts (84.4%), while less than 3% were not rostered in each cohort.

Comorbidity burden was substantial in both cohorts: only about 10% had no recorded chronic conditions in addition to cancer, while nearly one in five had five or more comorbidities.

Cancer stage at diagnosis had substantial missingness (approximately 50%), limiting interpretation of stage-specific differences. Nevertheless, most cancer diagnosed were at stage 1 in both cohorts, with a marked differences in stage 4 cancer in the pandemic cohort (7.2%) compared to the pre-pandemic cohort (10.4%). The most frequent cancer sites were similar in the pre-pandemic cohort (breast 14.6%, prostate 12.4%, and colorectal 9.8%) and the pandemic cohort (breast 16.6%, prostate 13.8%, and colorectal 9.8%), with however slightly less lung cancers in the pandemic cohort (8.7%) compared to the pre-pandemic cohort (10.9%).

Compared to the general Ontario population without cancer, patients in the cancer cohorts were substantially older and had markedly higher comorbidity burden (Supplemental Table 1). Detailed subgroup analyses for all outcomes are provided in Supplemental Tables 2–18, with case definitions in Supplemental Tables 19–21.

## Key Findings Organized by Themes

### Theme 1: Patients Experienced Significant Delays with Cancer Diagnosis and Screening

Participants consistently described major disruptions across the cancer diagnostic pathway, including screening, imaging, pathology, and referral processes. Kimber (manager) described how public health messaging discouraged emergency department use, while primary care offices were often not seeing patients in person:

> “I’ve never seen such sick patients. Because they didn’t have their colonoscopies. They didn’t have their pap smears. And they didn’t have their mammograms. For two years, screening stopped. … Patients were told through the media and public, “don’t come to emerge unless it’s an emergency. Only see your family physician for non-emergent things.” And family physicians were like, “we’re not seeing anyone.” … So, patients were not accessing healthcare at all really for two years. And so, their symptoms became worse and worse and worse until it got to the point where it became an emergency. … And by the time they come to us now on inpatients they have such advanced disease.”

Participants also described the suspension of public health reminders. Jackie (family physician) explained:

> “When those letters stopped, people forgot that they needed screening.”

Fear of infection further discouraged care-seeking. Lex (nurse) noted that people “didn’t want to go to the emergency department [because] they were afraid of COVID,” and Jackie (family physician) emphasized that “the fear was huge.”

#### Screening and emergency department use

Quantitative analyses partially aligned with these accounts. Cervical and breast cancer screening rates declined during the pandemic. The proportion of eligible patients receiving cervical screening decreased from 64.8% to 57.5% (p=0.001), and breast screening declined from 64.5% to 57.2% (p=0.008) (Figures 3 and 4; Supplemental Tables 3–4). In contrast, colorectal screening remained stable overall (73.3% pre-pandemic vs. 73.2% during the pandemic (Figure 2; Supplemental Table 2)

**Figure 2:**
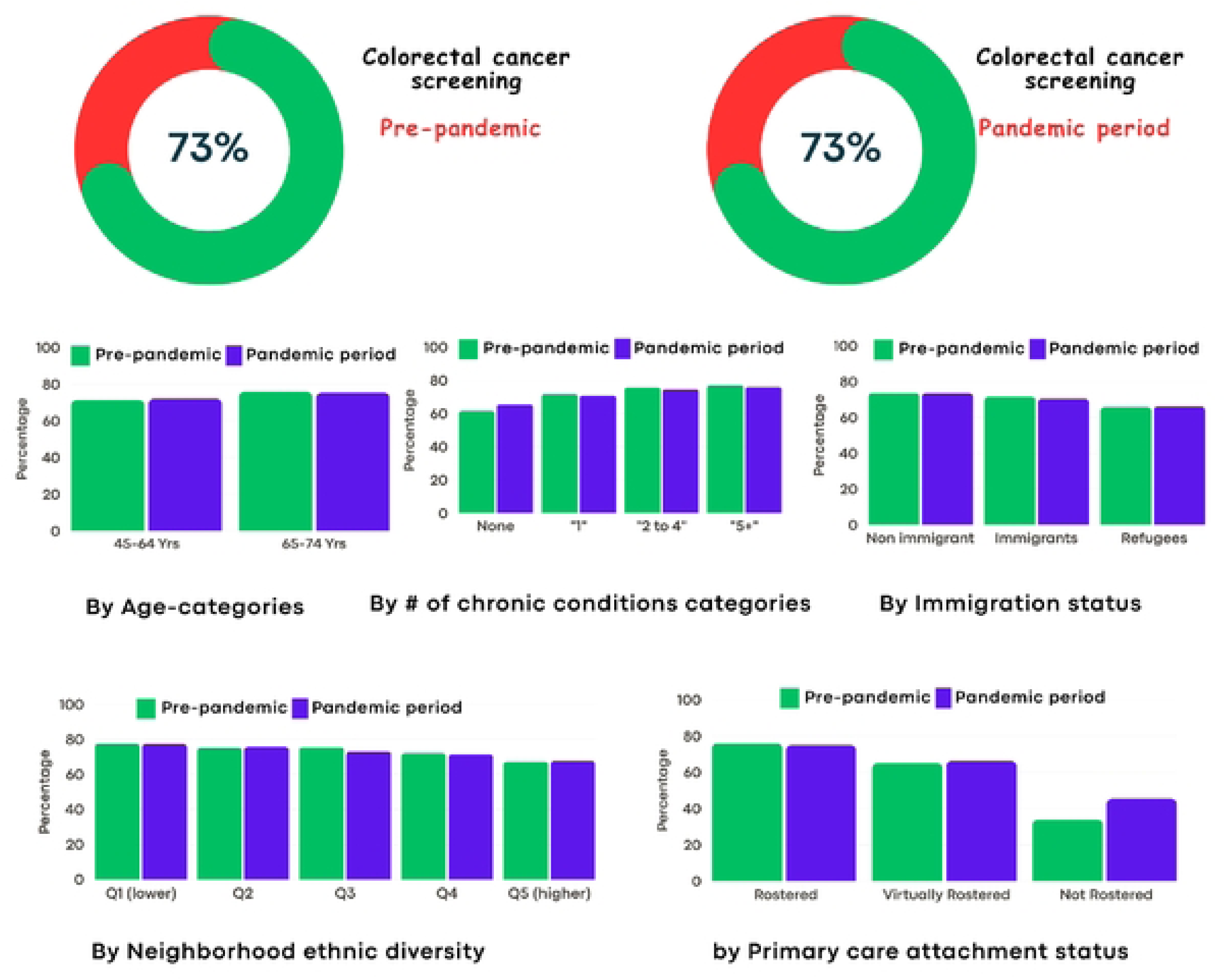
Percentage of eligible cancer patients who had colorectal cancer screening

**Figure 3:**
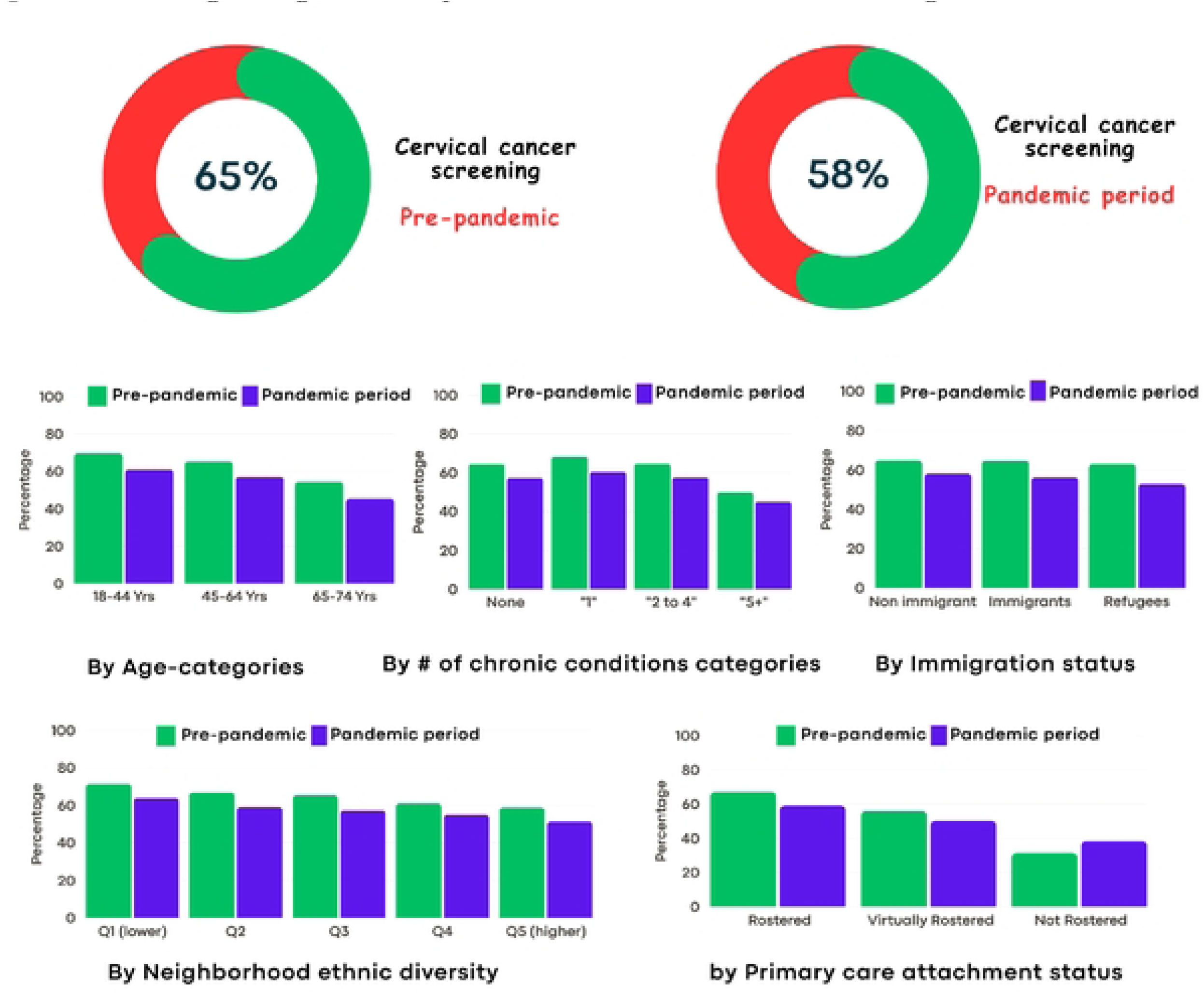
Percentage of eligible cancer patients who had cervical cancer screening

**Figure 4:**
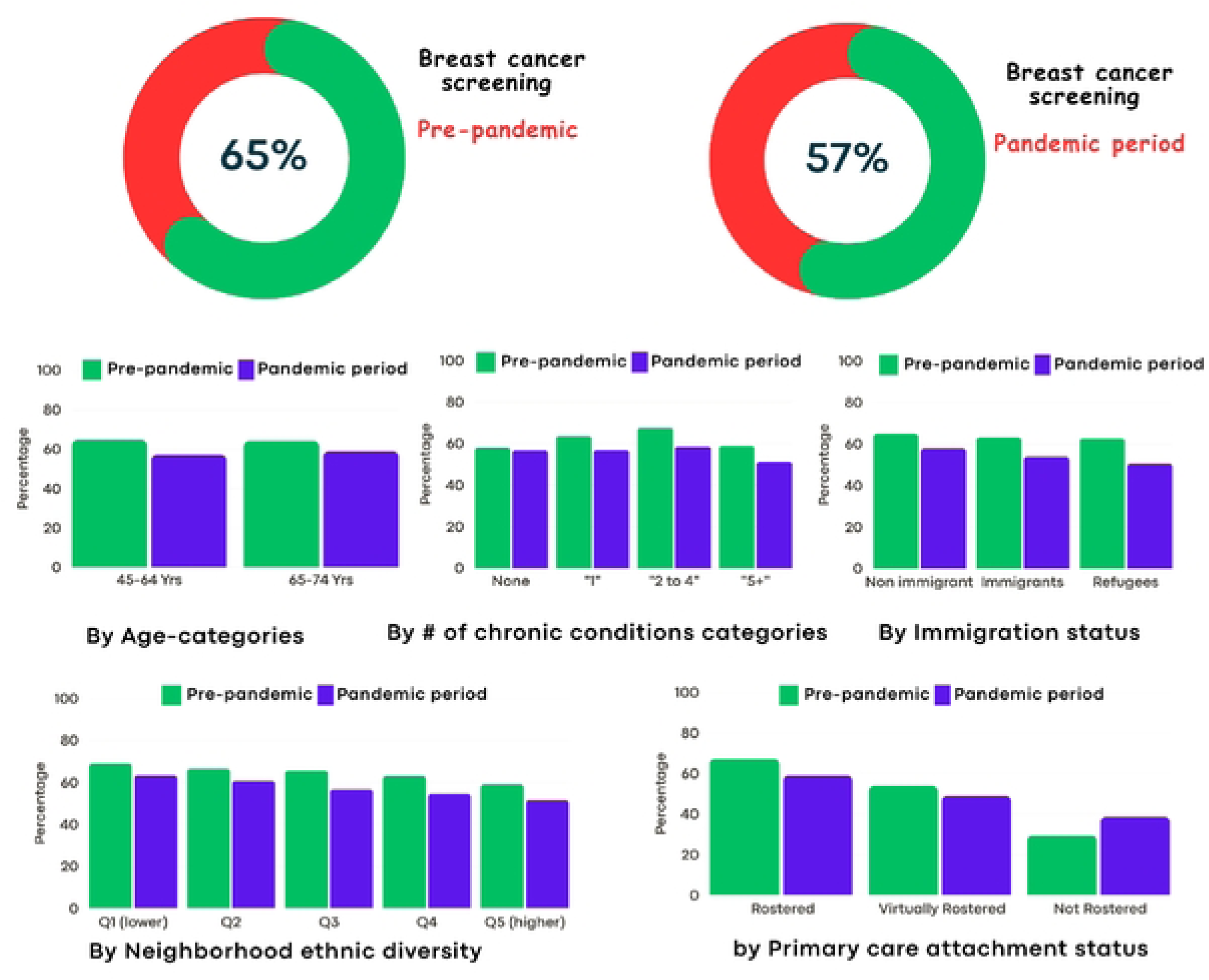
Percentage of eligible cancer patients who had breast cane.er screening

Reductions in cervical and breast screening were observed across most demographic and clinical subgroups. Declines were evident across neighbourhood ethnic diversity quintiles and immigration categories, including refugees. Patients rostered to a primary care provider had consistently higher screening participation than those not rostered, both before and during the pandemic. Notably, non-rostered patients had substantially lower screening rates overall (e.g., colorectal screening 34.2% pre-pandemic vs. 45.3% during the pandemic), indicating persistent structural differences in access to screenings.

Emergency department (ED) utilization also declined during the pandemic. The proportion of cancer patients with at least one ED visit decreased from 59.0% to 53.4% (p=0.002) (Figure 5; Supplemental Table 5). This reduction was observed across age groups, rural and urban settings, and multimorbidity levels. Patients with five or more comorbidities continued to have the highest ED utilization in both periods (81.1% pre-pandemic vs. 73.2% during the pandemic), although rates declined during the pandemic. Avoidable ED visits were uncommon overall for these cancer patients (<2% in both periods) but decreased by approximately half during the pandemic (1.6% to 0.8%, p=0.004; Supplemental Table 7).

**Figure 5:**
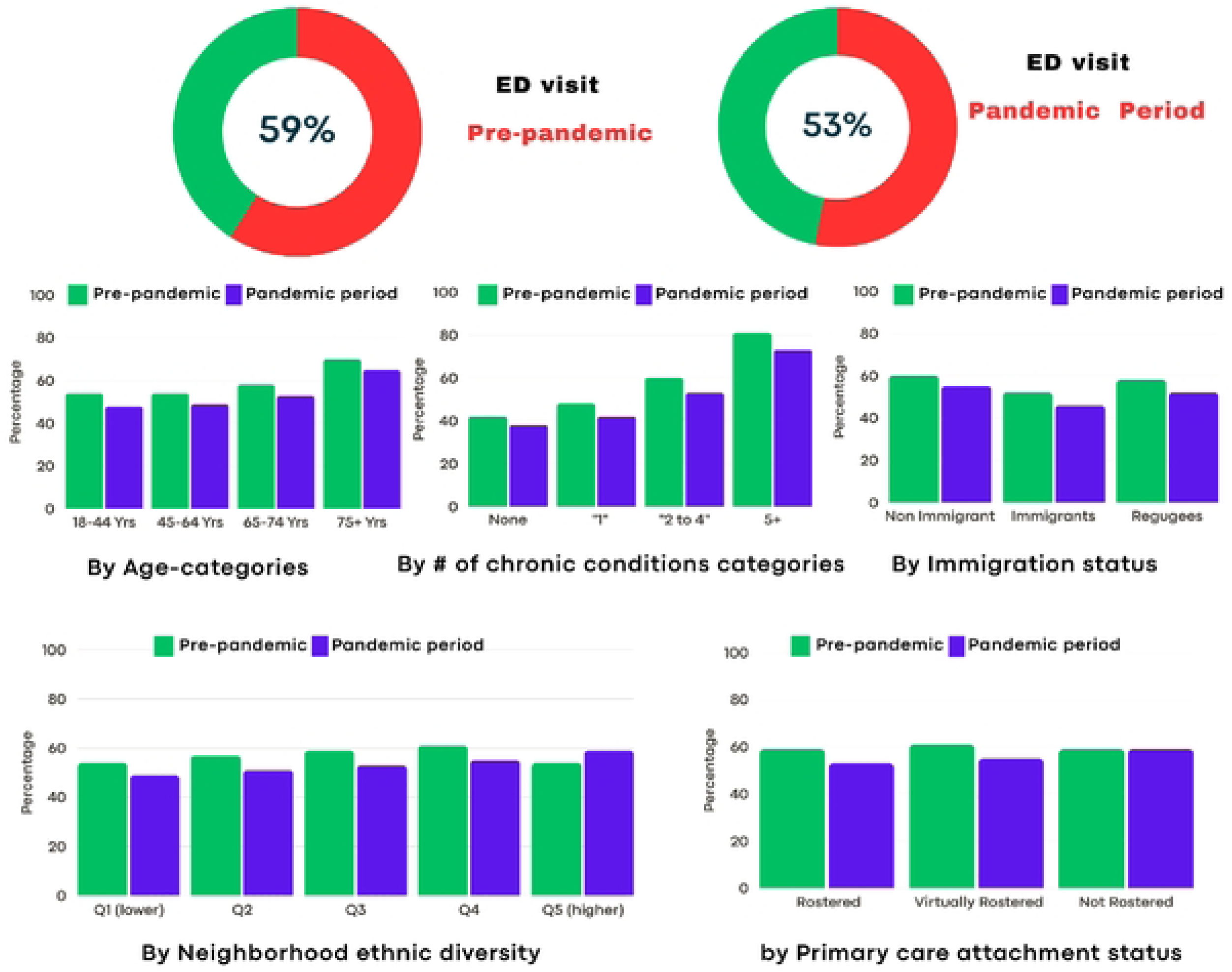
Percentage of cancer patients who had at least one emergency department (ED) visit

#### Diagnostic testing delays

Beyond screening and ED use, qualitative participants described substantial delays in imaging and diagnostic procedures. Jules (caregiver/clinical educator) stated that “the biggest delay was really getting the diagnosis.” Macey (caregiver/nurse) described delays in CT imaging:

> “In terms of diagnostic tests, I will say a number of my phone calls are patients frustrated that their CT scans are delayed. When the doctor told them it was going to be done in two weeks and it’s not been even booked yet. And it’s now almost [been] a month. … Overall, on the whole spectrum, patients are missing out on something.”

She recounted another case in which an MRI requested within 10 days was not booked for over three months:

> “The radiation oncologist wanted an MRI of her liver, and she asked for it in 10 days … and it wasn’t booked. … That doesn’t seem reasonable. Like you asked for it in 10 days … but it shouldn’t be three and a half months.”

Bronaugh (patient) described a prolonged interval between symptom recognition and biopsy:

> “I ended up getting my mammogram and my ultrasound in the middle of November. So that was two months later [after seeking primary care]. … Another month goes by. Pretty much December [date] of 2020 is when I get my biopsy for the lumps. … I discovered the lump in February of 2020, by the time I got in for a biopsy, it was end of December of that year. So, it was a good 10, 11 months before I got in.”

Participants also described lost requisitions and communication breakdowns. May (patient) explained:

> “I kept asking in my appointments. I was, like, “I haven’t heard about this brain skull scan”. And they kept saying, “Oh, we put the order in. It’s coming, it’s coming.” And I finally was like, “something’s wrong.” So, I called the diagnostic imaging centre … and they’re like, “We don’t have any requisition.” … In my experience, the scanning completely fell apart. And I don’t know. I don’t know whether it’s just more patients, or less staff, or both.”

Nidda (clinical manager) attributed some of these delays to staffing shortages:

> “They’re short staffed because there’s more people leaving the profession than coming in. There’s a delay in those pathology reports and so a delay in getting access to cancer service”.

#### Synthesis

Overall, qualitative participants described significant disruptions to screening and diagnostic pathways. Quantitative data corroborated declines in cervical and breast screening and reduced ED utilization during the pandemic, while colorectal screening remained stable. It was not possible to investigate the diagnostic testing delays in the quantitative data. However, together, these findings indicate measurable reductions in preventive and acute care utilization during the pandemic period, alongside qualitative accounts of delayed diagnostic processes and increased clinical severity at presentation.

### Theme 2: Difficulty Accessing Healthcare Through Primary Care

#### Perceived disruptions in continuity and access to primary care

Participants described primary care as a critical gateway to cancer diagnosis and system navigation. Jackie (family physician) emphasized that:

> “Primary care [was] one of the most stressful and deepest areas to affect cancer care because we are at the diagnostic level.”

She described primary care as:

> “a messy arena of trying to pick up stuff, trying to navigate specialists, trying to navigate diagnostic imaging, and advocate for patients.”

However, during the pandemic, participants reported disruptions to in-person access. Bronaugh (patient) described how:

> “All the walk-in clinics, the regular ones, were shut down… everything got shut down, literally everything. Doctor’s offices.”

She continued:

> “There was no communication on what do you do [when you have a cancer symptom]. … My doctor’s office had no communication.”

Lex (nurse) similarly stated that “family physicians and stuff weren’t seeing patients.” Fear further limited access. Mani (discharge planner) reported hearing patients say:

> “‘I was just too scared to go to the doctor.’ Not scared because of what the doctor would say… but scared to actually leave the house because of COVID.”

Jackie reinforced that: “The fear was the biggest barrier.”

Participants also described frustration and anger related to perceived disengagement. Jules (caregiver/clinical educator) noted:

> “It turned into like anger because she wasn’t feeling that she was getting any answers…”

Danni (patient) described missed warning signs:

> “Every symptom I brought forward was dismissed… the person I trusted the most [family doctor] is the person I should have feared the most.”

Others highlighted competing demands on family physicians. Jules explained:

> “The family physician … was involved with setting up the COVID screening clinic. So, I think she was being pulled in a lot of different directions at the same time.”

Together, these accounts describe perceived inaccessibility, communication breakdown, and diminished continuity of care.

#### Utilization of care and its modality

In contrast to qualitative reports suggesting reduced primary care access, administrative data indicated that overall physician visit volume per patient remained relatively stable. The median number of physician visits declined slightly from 26 (IQR 12–50) pre-pandemic to 24 (IQR 11–49) during the pandemic (Supplemental Table 8). Although statistically significant, this difference was modest in magnitude.

The key change was not overall volume, but modality. The proportion of patients receiving virtual (non-face-to-face) primary care increased dramatically from 8.5% pre-pandemic to 95.4% during the pandemic (p < 0.001; Supplemental Table 9 and Figure 6). This shift was observed across age groups, rural and urban settings, immigration categories, and multimorbidity levels. Virtual care uptake was slightly lower among patients residing in neighbourhoods with the highest ethnic diversity quintile (90.5%) and among patients not rostered to a primary care provider (88.9%), though levels remained high overall.

**Figure 6:**
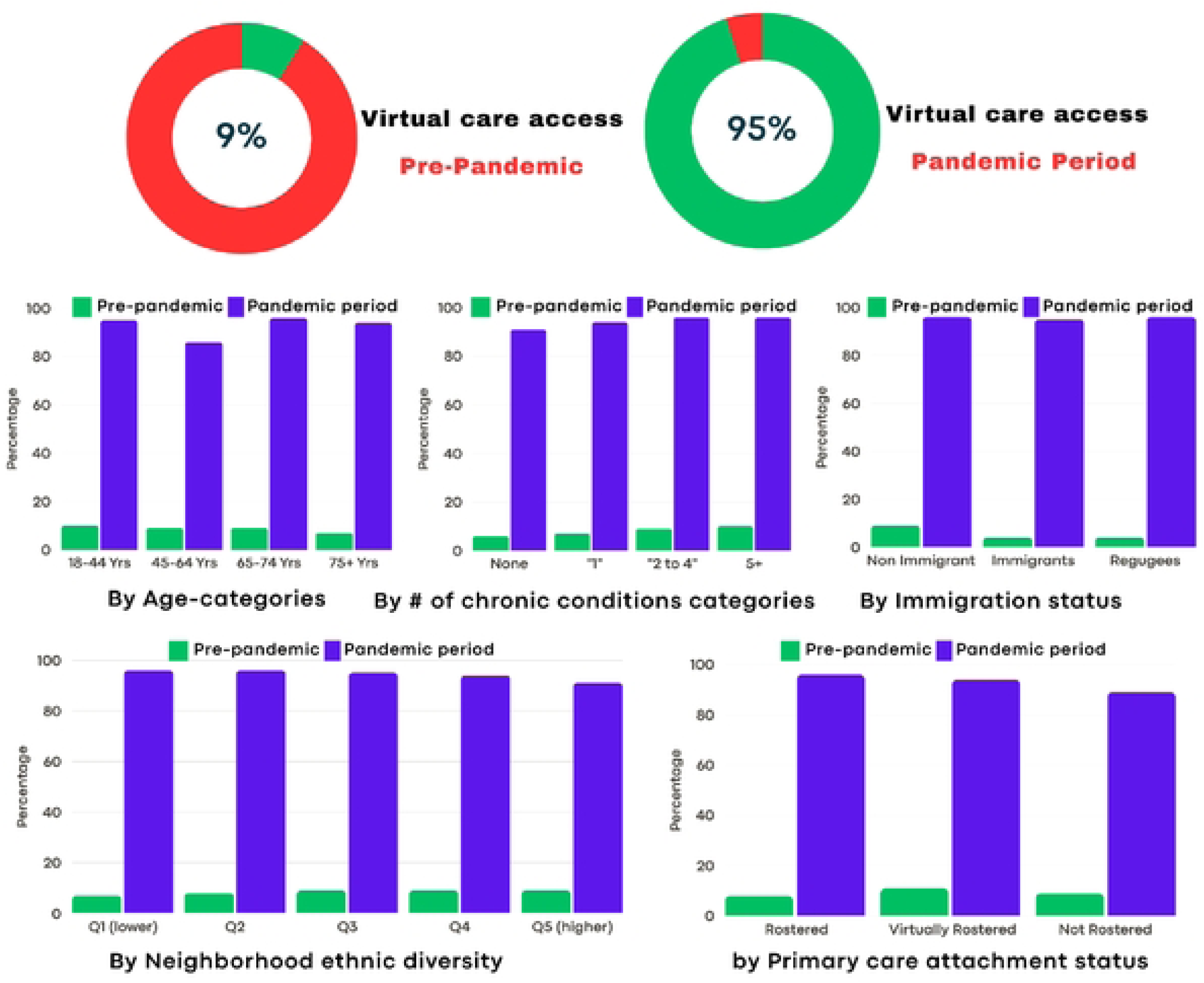
Percentage of cancer patients who received virtual care

The proportion of patients virtually rostered to a primary care provider remained stable (13.5% pre-pandemic vs. 13.1% during the pandemic; Supplemental Table 10). Refugees and immigrants had higher rates of virtual rostering compared to non-immigrants in both periods.

#### Reconciling qualitative and quantitative findings

The apparent tension between qualitative accounts of limited access and quantitative evidence of stable visit volume can be understood by distinguishing between utilization and experiential access.

While most patients had at least one contact with primary care during the pandemic, qualitative participants described substantial barriers in navigating telephone-based care, prolonged wait times, and reduced perceived responsiveness. Jules explained:

> “Sometimes it would be over an hour to get through to the family doctor… I think it was just the access.”

Thus, although the administrative data capture contact with providers, they do not reflect delays in reaching providers, communication challenges, or perceived quality of interaction.

No quantitative measures were available to capture fear-driven avoidance, competing provider responsibilities, or dissatisfaction with care processes. Administrative data record utilization, but not unsuccessful attempts to obtain care or patient perceptions of access.

#### Synthesis

Qualitative findings highlighted disrupted continuity, communication barriers, and fear-based avoidance in primary care. Quantitative data demonstrated that overall physician visit volume remained relatively stable, but care delivery shifted almost entirely to virtual modalities. Together, these findings suggest that while formal contact with primary care was largely maintained, the mode and experience of access changed substantially during the pandemic.

### Theme 3: Difficulty Accessing Specialists

#### Perceived reductions and delays in specialist access

Qualitative participants described substantial challenges in accessing specialist care during the pandemic. Referrals were delayed, lost, or returned without timely appointments. Mateo (patient) described a referral for a liver cancer lesion:

> “A referral was made to [location] to have the lesion removed. Never heard back. … That was a considerable delay.”

Jackie (family physician) characterized referral processes as:

> “Terrible. Just terrible. People just weren’t seeing patients. … Unless you were emergent, you wouldn’t be seen.”

She explained that specialists reduced in-person capacity due to infection control requirements, and further described receiving delayed booking notices or assuming dermatologic procedures typically managed by specialists:

> “The specialists cut back on the number of patients they could see because they used to have very crowded waiting rooms.”
>
> “Your referral is now eighteen months away.”
>
> “There’s no one who will do that. It’s now us.”

West (nurse) noted that referrals were sometimes misdirected:

> “They should’ve been referred to us, but they were referred to the wrong [specialist] area. Because their care was delayed, their cancer was diagnosed later.”

Participants described family physicians absorbing additional workload when specialist access was constrained. Macey (caregiver/nurse) recounted leveraging personal connections:

> “Even though we didn’t actually have a biopsy… he was willing to talk to her because we had no one else to talk to.”

Jackie (family physician) emphasized:

> “I can’t even begin to start with the burden that is landed on primary care.”

Together, these accounts reflect perceived contraction of specialist capacity and increased burden on primary care.

#### Variations across specialties

Administrative data revealed a more differentiated pattern.

The proportion of cancer patients with at least one oncology follow-up increased during the pandemic (45.0% pre-pandemic vs. 55.6% pandemic, p<0.001; Supplemental Table 11 and Figure 7). This increase was observed across age groups, sex, rural and urban settings, immigration categories, and comorbidity levels. The largest absolute increases were seen among adults aged 45–74 and among patients not rostered to primary care (42.6% to 63.6%). However, the median number of oncology visits per patient declined slightly (7 [IQR 3–18] pre-pandemic vs. 6 [3–17] pandemic; Supplemental Table 12), suggesting that while a greater proportion of patients had at least one oncology follow-up, visit intensity per patient did not increase.

**Figure 7:**
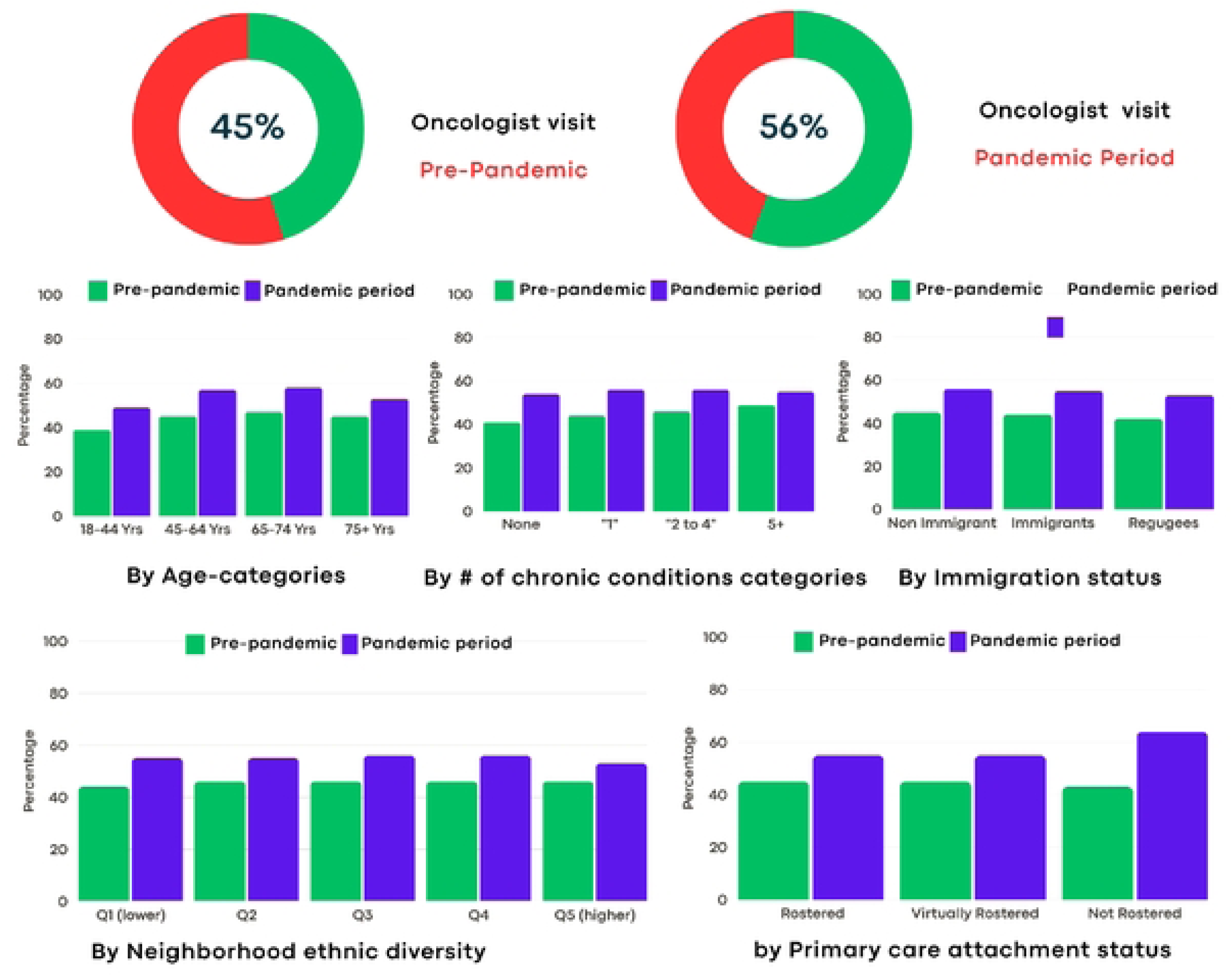
Percentage of cancer patients who had at least one follow-up by an oncologist

In contrast, non-oncology specialist visits declined. The proportion of patients with at least one cardiology visit decreased from 55.0% to 48.7% (p<0.001; Supplemental Table 13 and Figure 8). Declines were observed across age groups and were most pronounced among patients with higher multimorbidity (80.6% to 72.9% among those with ≥5 comorbidities). Similarly, although absolute rates were low, outpatient mental health visits decreased modestly (0.4% to 0.3%, p=0.009; Supplemental Table 15).

**Figure 8:**
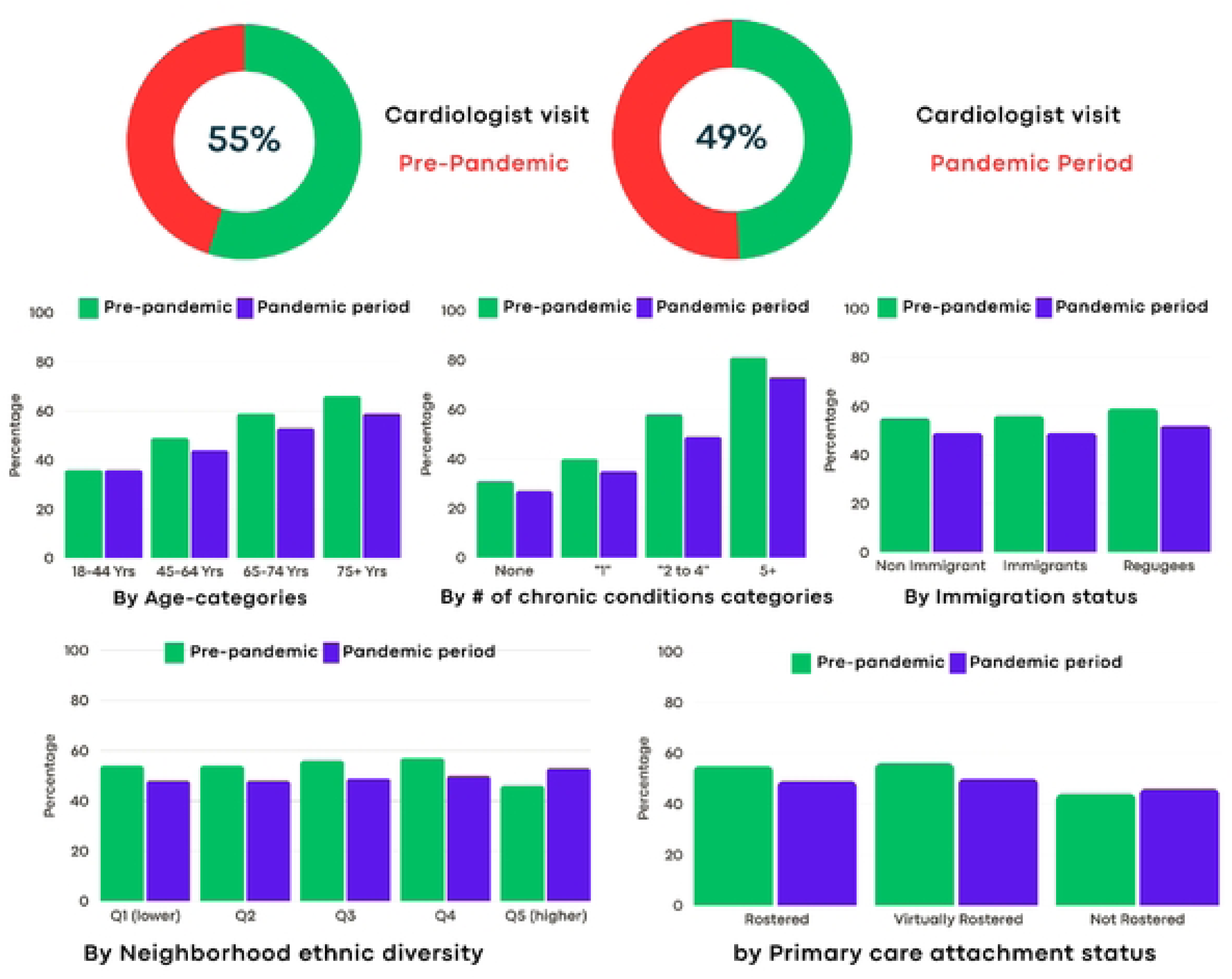
Percentage of cancer patients who have ever received a cardiologist visit

Lastly, endocrinology visits remained relatively stable (11.2% to 11.8%; Supplemental Table 16), with minimal changes in visit intensity (Supplemental Table 17).

Taken together, the quantitative findings do not suggest a uniform reduction in specialist access. They indicate differentiated effects across specialty types. Oncology follow-up was maintained or expanded in terms of the proportion of patients seen, but with lower intensity, while cardiology and mental health services declined.

#### Synthesis

Administrative data do not capture referral attempts, delays prior to first appointment, or time to consultation, contrary to the qualitative findings. Thus, while oncology follow-up rates increased, qualitative accounts of delayed or restricted specialist access likely reflect barriers in referral processes, booking timelines, or perceived responsiveness rather than complete absence of specialist contact. Together, these findings suggest that specialist access during the pandemic was not uniformly reduced but instead shifted in ways that prioritized oncology care in presence of constrained access to other specialties.

### Theme 4: Fragmented Care for Patients with Complex Needs and Chronic Conditions

#### Navigating siloed care across multiple conditions

Participants described the heightened complexity of managing cancer alongside multiple chronic conditions during the pandemic. Mani (discharge planner) noted that:

> “People are coming in with more than one [condition], more than just the cancer.”

Beth (patient) described her care as:

> “Basically disjointed… there [was] no one to look at you totally to see what’s wrong.”

May (patient) illustrated the siloed structure of care:

> “You don’t ask your plumber what’s wrong with your car. And you don’t ask your mechanic to fix your lighting electrical issues. And oncology’s no different. … It’s just a matter of finding out, when I’ve got questions, who runs that silo.”

Beth further described poor coordination across specialties:

> “I phoned the cancer clinic to ask, ‘should I be taking these medications when I’m getting all this other medication for the pericarditis?’ They didn’t phone me back for a week.”

Participants emphasized that patients often had to assume responsibility for coordinating information across providers and ensuring medication safety.

Ingrid (nurse) observed that patients prioritized cancer treatment over management of other chronic conditions:

> “Patients were a little less likely to go to other appointments for other chronic issues… they will pretty much put their cancer treatment at the biggest priority.”

She further noted that additional diagnostic requirements for comorbid conditions (e.g., cardiac testing prior to chemotherapy) sometimes resulted in delays:

> “Often, patients just weren’t really going through with that. So that was a little bit of a delay.”

Fear of COVID exposure compounded this:

> “Often, patients with other chronic illnesses… would just not really want to go and do those [tests] just to not get exposed to COVID.”

Ingrid concluded:

> “We’re so hyper focused on one thing… those chronic conditions are getting worse, worse, worse, and maybe not noticed until a certain point.”

Together, these accounts describe fragmentation across care silos, increased patient burden, and deprioritization of non-cancer conditions.

#### Patterns by multimorbidity

Administrative data showed consistent differences in healthcare utilization by comorbidity burden.

Patients with higher multimorbidity had substantially higher emergency department (ED) use in both periods. Among patients with five or more comorbidities, ED use declined from 81.1% pre-pandemic to 73.2% during the pandemic (Supplemental Table 5 and Figure 5) yet remained markedly higher than among patients without comorbidities.

Screening patterns also varied by comorbidity level. For cervical and breast cancer screening, declines were observed across comorbidity strata, with larger absolute reductions among patients with moderate to high comorbidity (Supplemental Tables 3–4 and Figures 3-4). In contrast, colorectal screening remained relatively stable across comorbidity levels (Supplemental Table 2 and Figure 2).

Non-oncology specialist care showed differential effects by comorbidity. The proportion of patients with five or more comorbidities who had at least one cardiology visit decreased from 80.6% pre-pandemic to 72.9% during the pandemic (Supplemental Table 13 and Figure 8). While oncology follow-up increased across comorbidity levels, the relative difference with the pre-pandemic became gradually less large as the number of comorbidities rose (Supplemental Table 11 and Figure 7).

#### Integrating qualitative and quantitative findings

Quantitative data demonstrate that patients with multimorbidity consistently had higher healthcare utilization, particularly emergency and specialist services. During the pandemic, oncology follow-up was maintained or increased across comorbidity levels, while cardiology visits and screening participation declined.

These patterns align with qualitative accounts suggesting prioritization of cancer care over other chronic conditions and increased fragmentation across specialty silos. While administrative data cannot capture care coordination challenges, delays in requisition completion, or patient perceptions of burden, the observed decline in non-oncologic specialist visits among highly comorbid patients supports the interpretation that management of co-existing conditions was comparatively constrained during the pandemic.

#### Synthesis

Patients with multiple chronic conditions experienced greater baseline healthcare utilization and greater complexity in navigating care. During the pandemic, oncology care was largely preserved, but non-oncologic and preventive services declined across comorbidity levels.

Together, qualitative and quantitative findings suggest that pandemic-related disruptions may have intensified fragmentation for patients with complex health needs, reinforcing siloed patterns of care.

## Discussion

### Summary of Principal Findings and Contribution

This sequential mixed-methods study examined how the COVID-19 pandemic affected cancer care accessibility in Ontario by integrating qualitative experiences with population-level administrative data. Four themes emerged from qualitative analyses: (1) delays in diagnosis and screening, (2) difficulty accessing primary care, (3) difficulty accessing specialists, and (4) increased challenges for patients with multimorbidity. Quantitative indicators were selected to assess whether these themes were observable at the population level. Overall, the qualitative themes were largely reflected in administrative data, while also revealing important differentiation across care domains and patient subgroups.

Both data sources indicated that disruptions to cancer screening and diagnostic pathways were among the most significant impacts of the pandemic. Cervical and breast cancer screening declined during the pandemic, whereas colorectal screening remained stable. Qualitative accounts contextualized these findings, describing suspended reminder systems, clinic closures, emergency department restrictions, and fear of COVID-19 exposure. The stability of colorectal screening may reflect the at-home nature of first-line stool-based testing, which reduced reliance on in-person care. These findings align with systematic reviews and population-based studies reporting reductions in cervical and breast cancer screening during the pandemic (4, 42, 43).

Reductions in screening were observed across multiple subgroups, including immigrants, refugees, and residents of high ethnic diversity neighbourhoods. Patients not rostered to a primary care provider had consistently lower screening participation in both periods, suggesting persistent structural differences in access rather than pandemic-specific effects alone. Prior research has similarly identified socioeconomic disadvantage, immigrant status, and limited primary care attachment as factors associated with lower cancer screening uptake (44–51).

Primary care access demonstrated a more complex pattern. Qualitative participants described inaccessibility, communication breakdown, and delayed referrals. In contrast, quantitative data showed that overall physician visit volume per patient remained relatively stable, but delivery shifted almost entirely to virtual modalities. This distinction highlights the difference between healthcare utilization and experiential access. Although administrative data indicated high levels of virtual contact, qualitative participants reported dissatisfaction, long wait times, and challenges navigating telephone-based systems. Similar rapid transitions to virtual care have been documented elsewhere, with variation in uptake across marginalized populations (52, 53). Together, these findings suggest that while formal contact with primary care was maintained, the experience and continuity of access changed substantially.

Specialist access showed differentiated effects. Qualitative participants described delayed or restricted referrals, consistent with published reports of resource reallocation and suspended services early in the pandemic (54, 55). However, quantitative data indicated that the proportion of cancer patients receiving at least one oncology specialist follow-up increased during the pandemic, whereas cardiology and mental health visits declined. Other Ontario-based studies have reported similar patterns of maintained or improved access to certain cancer-directed services during the pandemic (20). These divergent patterns may reflect simultaneous organizational processes: early restrictions that delayed diagnosis and referral, alongside prioritization of ongoing cancer treatment for patients already connected to oncology services.

Finally, both qualitative and quantitative findings highlighted challenges for patients with multimorbidity. Participants described fragmented care across specialties and increased burden navigating siloed systems. Quantitatively, patients with higher comorbidity burden had consistently higher healthcare utilization, including emergency department use and specialist care. These findings suggest that cancer-directed care may have been prioritized, while management of co-existing conditions experienced comparatively greater disruption. Previous studies have shown that multimorbidity complicates cancer care and is associated with poorer outcomes (56, 57); the pandemic context may have intensified these coordination challenges.

Taken together, this study contributes to the literature by integrating lived experiences with population-level utilization data within a single analytic framework. Rather than examining qualitative or quantitative evidence in isolation, this approach enabled identification of both concordance and divergence between experiential and system-level patterns. The findings indicate that pandemic-related disruptions were not uniform across care domains, but varied by modality, specialty type, and patient characteristics.

### Strengths and Limitations

A key strength of this study is its sequential mixed-methods design, which enabled integration of experiential qualitative findings with population-level administrative data (23, 24, 58). The qualitative phase identified barriers and disruptions in cancer care from the perspectives of patients, caregivers, clinicians, and system leaders. The subsequent quantitative phase assessed whether these patterns were observable at scale using linked administrative databases. This approach supports triangulation across data sources while allowing identification of both concordance and divergence between lived experience and system-level utilization patterns (23, 24, 58).

The use of comprehensive provincial administrative databases minimized recall bias and enabled population-level inference, including subgroup analyses across immigration status, neighbourhood ethnic diversity, multimorbidity burden, and primary care enrolment. At the same time, qualitative interviews provided contextual insight into referral processes, communication challenges, and perceived barriers that are not captured in utilization data.

Several limitations should be considered. First, the qualitative component included a modest number of patients and caregivers. Although participants represented multiple roles within the cancer care system, additional patient perspectives may have yielded further nuance. Second, administrative data capture healthcare utilization but do not measure unsuccessful attempts to obtain care, perceived access, quality of interaction, or clinical outcomes beyond service use. Thus, some experiences described in the qualitative findings (e.g., difficulty reaching providers, dissatisfaction with virtual interactions) cannot be directly assessed using administrative indicators. Third, comorbidity was examined using aggregate counts of chronic conditions. While this enabled consistent subgroup analyses, it may not capture condition-specific barriers experienced by patients with particular chronic illnesses. Fourth, the study did not examine differences by cancer site in depth. Prior Ontario studies using administrative data have reported variation in screening, new diagnoses, and specialist follow-up by cancer type when comparing pre-pandemic and pandemic periods, suggesting the value of future site-specific analyses (19, 21, 59). In addition, stage at diagnosis had substantial missingness in administrative data, limiting usefulness of the data to interpret stage-specific differences. Fifth, the quantitative analyses were based on administrative data available through December 2021, corresponding to the data cut available when qualitative data collection commenced. While this allowed alignment of the two phases within the study timeline, longer-term effects beyond this period were not assessed. Finally, mixed-method integration remains methodologically complex, and limited guidance exists on best practices for integrating qualitative findings with health administrative data (23, 24, 58). Differences observed between experiential accounts and utilization patterns may therefore reflect measurement limitations.

### Implications for Policy and Research

The findings of this study suggest that pandemic-related disruptions in cancer care were not uniform but varied across care domains and patient groups. Screening and diagnostic pathways were particularly vulnerable, while oncology follow-up was largely preserved. These differentiated patterns indicate that healthcare systems may prioritize active cancer treatment during crises, but preventive services and non-oncologic specialty care may experience greater disruption.

From a policy perspective, safeguarding access to timely cancer screening and diagnostic services during public health emergencies is critical. The observed declines in cervical and breast cancer screening, particularly among immigrants, refugees, and patients not rostered to primary care, underscore the importance of maintaining outreach, reminder systems, and primary care attachment for structurally vulnerable populations (44–48). Strategies that protect preventive services alongside acute and specialist care may reduce downstream delays in diagnosis and treatment.

The rapid transition to virtual primary care preserved contact with providers but did not eliminate perceived barriers. Future policy efforts should consider not only availability of virtual services, but also navigation supports, continuity mechanisms, and equitable access across population groups (52, 53). Ensuring that primary care remains a stable point of entry into the healthcare system may help mitigate delays in diagnosis and referral during future disruptions.

The findings also highlight the need for improved coordination for patients with multimorbidity. Although oncology services were maintained, cardiology and mental health visits declined, and qualitative accounts clearly suggesting potential fragmentation in the management of co-existing conditions. Strengthening integrated, team-based care models and enhancing communication across specialties may improve continuity for patients with complex health needs (56, 57).

From a research perspective, further work is needed to examine longer-term consequences of pandemic-related disruptions. Prior literature suggests that delays in screening and treatment may influence staging at diagnosis, healthcare utilization, and cancer outcomes in subsequent years (60–62). Longitudinal analyses examining stage distribution, survival, and condition-specific outcomes will be important to assess the sustained impact of the pandemic. Future research may also explore cancer-site–specific differences and condition-specific multimorbidity patterns that could not be fully examined in the present study.

Finally, this study underscored the value of integrating qualitative and administrative data to better understand health system performance during crises (23, 24, 58). Continued methodological development in mixed-method integration may strengthen the ability to identify gaps between experiential access and utilization metrics in future system-level research.

## Conclusions

This mixed-methods study demonstrates that the COVID-19 pandemic was associated with measurable disruptions in cancer screening, diagnostic pathways, and selected specialist services in Ontario, while oncology follow-up was largely preserved. By integrating qualitative experiences with population-level administrative data, the study shows that system-level utilization patterns both reflected and qualified lived experiences of access barriers. Disruptions were not uniform, but varied by service type and patient characteristics, particularly for individuals with multimorbidity. Strengthening preventive service continuity, primary care coordination, and integrated management of complex conditions may enhance the resilience of cancer care systems during future public health emergencies.

## Data Availability

For qualitative data, our small sample and specific ethics board approvals in two Ontario hospitals make the respondents potentially identifiable. The data contains sensitive patient information and cannot be broadly shared. Contact the corresponding author who will then follow-up with the two research ethics boards to consider request for de-identified data access on a case-by-case basis. For the quantitative data, the administrative data is owned by a third-party, ICES. Specific processes are in place to access the data for public and private researchers, and are explained at the following links: https://www.ices.on.ca/services-for-researchers/public-sector-researchers/ and https://www.ices.on.ca/services-for-researchers/private-sector-researchers/

## Acknowledgement

We thank the participants of the qualitative portion of the study, who dedicated their time and energy to sharing their experiences and perspectives. We also acknowledge the broader research team that informed the work in the early stages. Dr. Kuluski holds the Dr. Mathias Gysler Research Chair in Patient and Family Centred Care, and Dr. Wodchis holds a Research Chair in Implementation and Evaluation Sciences both funded by the Trillium Health Partners Foundation.

## ICES statement

This study contracted ICES data & Analytic Services (DAS) and used de-identified data from the ICES Data Repository, which is managed by ICES with support from its funders and partners: Canada’s Strategy for Patient-Oriented Research (SPOR), the Ontario SPOR Support Unit, the Canadian Institute for Health Research, and the Government of Ontario. This study was supported through the provision of data by ICES and Cancer Care Ontario (CCO) and through Funding support to ICES from an annual grant by the Ministry of Health (MOH) and the Ontario Institute for Cancer Research (OICR). The opinions, results and conclusions reported in this paper are those of the authors. No endorsement by ICES or any of its funders or partners is intended or should be inferred.

## Funding

This work was supported by the Canadian Institutes of Health Research (CIHR) grant #179909. The funding source had no role in study design; in the collection, analysis, and interpretation of data; or in the writing of this manuscript.

## Notes

### Competing Interest Statement

The authors have declared no competing interest.

### Funding Statement

Yes

### Author Declarations

For the qualitative data portion, this study received Research Ethics Board approval from Trillium Health Partners (THP REB #1131) and Thunder Bay Regional Health Sciences Centre (TBRHSC REB #100217). All participants consented prior to engaging in the study by reading and signing an informed consent statement. For the quantitative data portion, the data come from health administrative databases held at ICES, Ontario and was authorized under section 45 of Ontario’s Personal Health Information Protection Act, which does not require review by a Research Ethics Board. All study investigations were performed in accordance with Canada’s Tri-Council Policy Statement: Ethical Conduct for Research Involving Humans.

